# Laser speckle contrast imaging for visualizing blood flow during cerebral aneurysm surgery: A comparison with indocyanine green angiography

**DOI:** 10.1101/2021.04.29.21254954

**Authors:** David R. Miller, Ramsey Ashour, Colin T. Sullender, Andrew K. Dunn

## Abstract

Laser speckle contrast imaging (LSCI) has emerged as a promising tool for intraoperative cerebral blood flow (CBF) monitoring because it produces real-time full-field blood flow maps non-invasively and label-free. In this study, we compare LSCI with indocyanine green angiography (ICGA) to assess CBF during aneurysm clipping surgery in humans. LSCI hardware was attached to the surgical microscope prior to the start of each surgery and did not interfere with the sterile draping of the microscope or normal operation of the microscope. LSCI and ICGA were performed simultaneously to visualize CBF in *n*=4 aneurysm clipping cases, and LSCI was performed throughout each surgery when the microscope was positioned over the patient. To more easily visualize CBF in real-time, LSCI images were overlaid on the built-in microscope white light camera images and displayed to the neurosurgeon in real-time. Blood flow changes before, during, and after an aneurysm clipping were visualized with LSCI and later verified with ICGA. LSCI was performed continuously throughout the aneurysm clipping process, providing the surgeon with immediate actionable information on the success of the clipping. The results demonstrate that LSCI and ICGA provide different, yet complementary information about vessel perfusion.

## Introduction

Cerebral blood flow (CBF) monitoring is crucial during cerebrovascular surgery to inform decision making.^1^ In cerebral aneurysm clipping cases, CBF monitoring is routinely used to confirm patency in vessels and determine successful aneurysmal obliteration. Current intraoperative tools for CBF monitoring and visualization include indocyanine green angiography (ICGA),^2-7^ Doppler^8-10^ and transit-time^11-12^ ultrasound, and percutaneous transfemoral digital subtraction angiography (DSA).^13-17^ ICGA records the fluorescence wash in of a bolus of indocyanine green after intravenous injection. DSA images are acquired by obtaining multiple time-controlled x-rays as contrast medium is injected intra-arterially.

During cerebrovascular procedures, it would be beneficial to assess CBF continuously, instead of a limited number of times^18^. ICGA is an effective decision making aid; however, it cannot provide continuous imaging as it requires an injected contrast agent. Doppler ultrasound provides absolute flow velocities, but is limited to measurement at single locations, and requires contact with the vessel of interest. DSA is the gold standard for confirming aneurysmal occlusion and patency of the underlying parent vasculature; however, it is invasive and time-consuming relative to ICGA or Doppler ultrasound, usually requiring removal of the surgical microscope, fluoroscopy, and transfemoral selective arterial catheterization.

Laser speckle contrast imaging (LSCI) has emerged as a promising tool to non-invasively monitor CBF because it produces real-time, full-field blood flow maps without any contrast agents, providing a potential continuous CBF monitoring solution. Both LSCI and ICGA images are restricted to the tissue surface, but their relatively simple instrumentation allows them to be incorporated into the surgical microscope. Several studies have demonstrated LSCI during neurosurgical procedures in humans and shown its promise as a CBF monitoring tool, including surgical revascularization,^19-21^ awake functional mapping,^22^ brain tumor resection,^23-25^ cortical spreading depression,^26^ and infarction during ischemic stroke.^27^ However, many previous clinical implementations required an external device to be introduced into the surgical field^27-29^ leading to disruptions of the surgical procedures. Other implementations incorporated the instrumentation into the neurosurgical operating microscope, eliminating the need for an external device and surgical disruption^23-25^. However, LSCI imaging still required the surgical procedure to be paused while images were acquired, and it was not possible to record CBF images for long durations, or simultaneously with ICGA.

In this article we demonstrate that LSCI integrated into the neurosurgical microscope allows real-time continuous visualization of CBF during neurosurgical procedures including simultaneous ICGA and LSCI imaging. We demonstrate these capabilities during cerebral aneurysm clipping surgery and compare LSCI directly with ICGA. Although simultaneous imaging of CBF with LSCI and ICGA has been performed in animal models,^30,31^ this comparison has not been performed in humans. This article demonstrates the potential of LSCI for human CBF monitoring in two ways: LSCI and ICGA were performed simultaneously to visualize CBF for *n*=4 aneurysm clipping cases, and LSCI was performed throughout each cerebral aneurysm clipping surgery without affecting the surgical work flow, including real-time visualization of CBF during aneurysm clip placement. Taken together, these results demonstrate that LSCI and ICGA provide different, yet complementary information about vessel perfusion.

## Methods

### Instrumentation

A schematic of the LSCI setup adapted to a surgical microscope (Leica M530 OH6, Leica Microsystems GmbH, Wetzlar, Germany) is shown in Fig. 1. The LSCI hardware (previously described in Richards et al.^24^) was attached to the microscope prior to the start of the surgery and did not interfere with the sterile draping or normal operation of the microscope. A λ=785 nm laser diode with a maximum output power of 300 mW was attached to an add-on laser adapter (MM6 Micromanipulator, Carl Zeiss Meditec Inc., Oberkochen, Germany). The laser adapter was mounted to the bottom of the microscope such that a steering mirror directed the light downward toward the surgeon’s field of view. The beam size was approximately 2 cm at a working distance of 35 cm. The maximum irradiance was 0.10 W/cm^2^, well below the American National Standards Institute (ANSI) limit of 0.3 W/cm^2^ for skin at 785 nm.^32^

**Figure 1.**
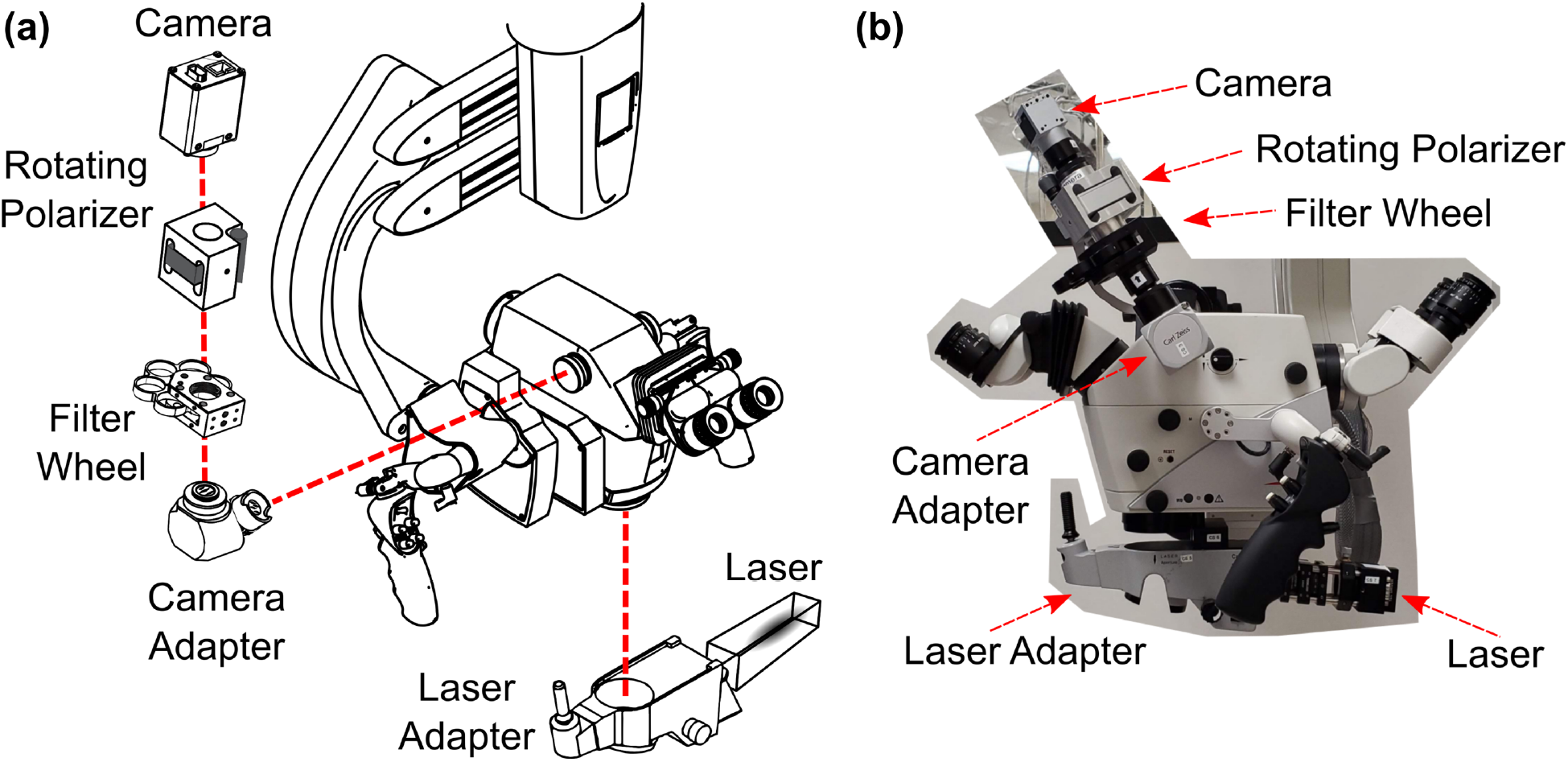
**(a)** Schematic of the Leica OH6 neurosurgical microscope outfitted with laser speckle contrast imaging (LSCI) instrumentation to intraoperatively measure cerebral blood flow. Drawings adapted from Leica M530 OH6 Brochure and Thorlabs Inc. filter wheel diagram. **(b)** Photograph of the LSCI hardware attached to the Leica OH6 microscope in the operating room.

Back-scattered laser light was directed to an NIR-enhanced CMOS camera (Basler AG, Ahrensburg, Germany) mounted on the side observer port on the same side as the craniotomy. This enabled an observer to participate during the study, which occurred for most of the surgery for patients 2 and 3, and intermittingly for patients 1 and 4. The pixel area was slightly cropped during acquisition to capture only pixels over brain tissue. A filter wheel (CFW6, Thorlabs Inc.) and polarizer (LPNIR100, Thorlabs Inc.) were positioned between the camera adapter and camera. The filter wheel held various neutral density filters for controlling the laser power. The polarizer was integrated into a motorized rotation mount (RSC–100, Pacific Laser Equipment Inc., Santa Ana, California, USA) to reduce specular reflections. A band-pass filter (FF01-788/3-25, Semrock Inc., Rochester, New York, USA) was added in front of the camera to enable simultaneous LSCI acquisition during illumination of indocyanine green and to block non-laser light and to avoid interference of normal white light illumination throughout each procedure.

### Image Analysis

ICGA images are represented as raw fluorescence intensity images that were collected by the built-in Leica OH6 NIR camera. Raw LSCI images collected by the speckle camera were processed by calculating the spatial speckle contrast value (*K*) within a 7 x 7 pixel moving window according to the equation 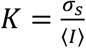, where σ_s_ is the spatial standard deviation and ⟨*I*⟩ is the average intensity within the region. For relative blood flow measurements, speckle contrast was converted into correlation time (τ_c_) by evaluating the average decay time of the speckle electric field autocorrelation function^33^ for which we assumed unity for the instrumentation factor. The speckle correlation time τ_c_ is a more quantitative measure of blood flow; the inverse correlation time (ICT=1/τ_c_) is commonly used as a metric for blood flow in vessels or perfusion in parenchyma.^34-37^ For displaying ICT time course data, the first five seconds of data was used as a normalization factor to more easily visualize the change in flow relative to the baseline value. To overlay LSCI with images from the built-in microscope white light camera, the video output of the surgical microscope system was recorded continuously. LSCI images, white light reflectance images, and ICGA images were spatially co-registered by applying an affine transformation to all corresponding images.

To better visualize CBF in real-time, LSCI images were thresholded and overlaid on the visible white light reflectance images, as illustrated in Fig. 2. First, the LSCI image is acquired, shown in Fig. 2A with a grayscale color map. Next, a threshold is applied to the LSCI image such that only speckle contrast values corresponding flow values within a certain range remain (i.e., high flow in vessels); additionally, a median filter and pseudo-color are applied for easier visualization, as shown in Fig. 2B. Following the threshold and pseudo-color, the LSCI image is merged with the white light image (Fig. 2C) along with desired transparency (Fig. 2D). Processing the LSCI frames, thresholding the high flow values, registering the thresholded LSCI image with the white light image, and overlaying the thresholded LSCI image onto the white light image to create the overlay image was performed with custom software on a desktop PC at video rate and displayed continuously to the neurosurgeon on a monitor in real-time.

**Figure 2.**
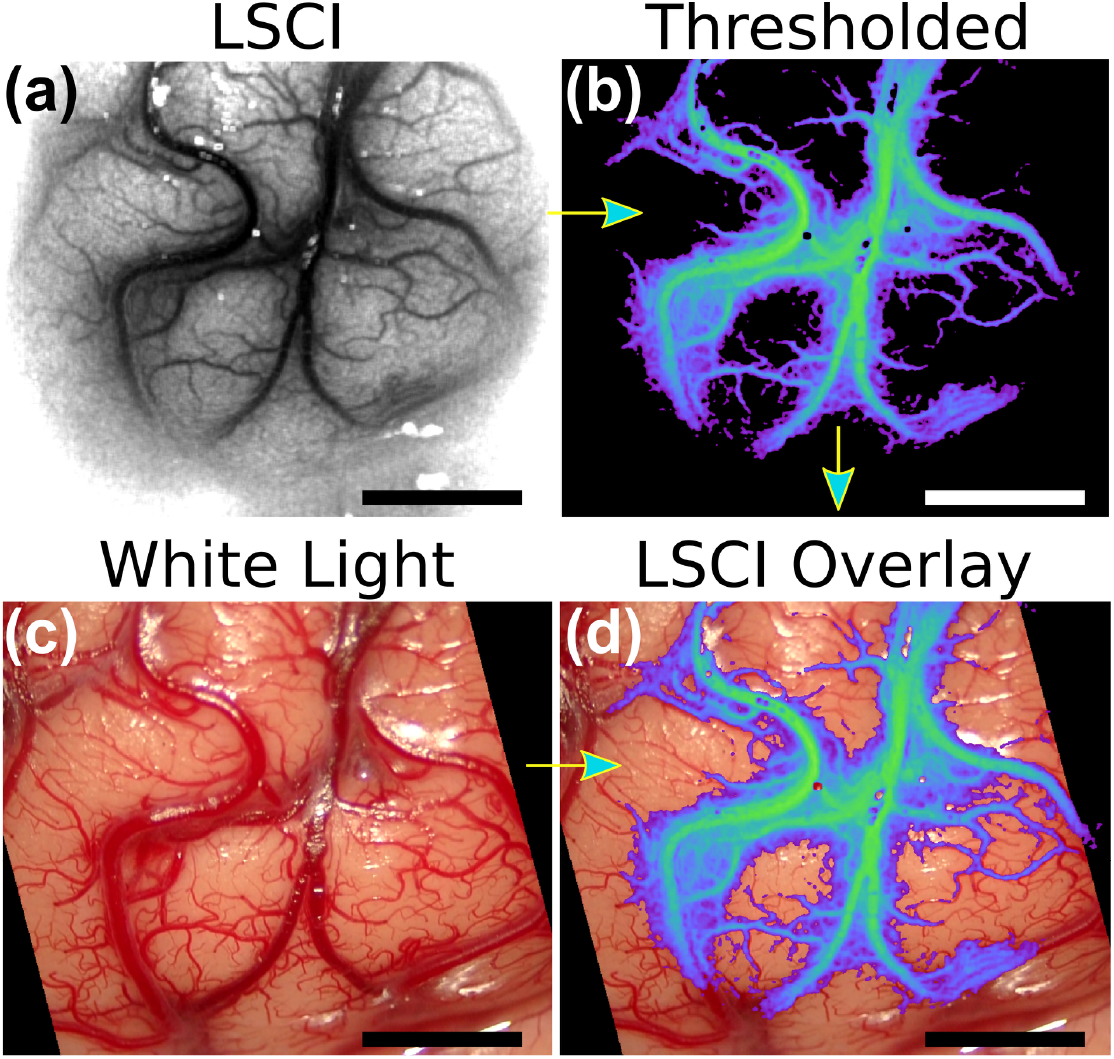
Image processing steps to create overlay of laser speckle contrast imaging (LSCI) on built-in microscope white light camera termed LSCI overlay. Data from patient 1 was used to create images. **(a)** LSCI image with grayscale color map. **(b)** LSCI image with a threshold applied to display high flow blood vessels, and median filter and pseudo-color applied. **(c)** White light image captured at the same time as LSCI image. **(d)** Thresholded pseudo-color LSCI image merged with the white light image to create an LSCI overlay image. Scale bars are 0.5 cm.

### Intraoperative Procedure

All *n*=4 surgeries were performed at Dell Seton Medical Center at the University of Texas at Austin by the neurosurgical co-investigator on this study (R.A.). The clinical study was approved by the Institutional Review Board of the University of Texas at Austin. Written and informed consent was obtained from all patients prior to surgery. A summary of patient details is shown in Table 1; more detailed patient information is located in the Supplemental Information.

**Table 1.** Patient details for each aneurysm case.

| Patient | Gender | Age Range | Aneurysm Location/Size | Preop Imaging | # times ICGA | Intraop and Postop Imaging | Clinical Outcome |
| --- | --- | --- | --- | --- | --- | --- | --- |
| 1 | F | 37-47 | Right SCA 5mm | CTA DSA | 1 | Intraop DSA Postop CTA | Transient CN III palsy resolved |
| 2 | F | 59-69 | Right ICA 11mm | CTA DSA | 2 | Intraop DSA | No complication |
| 3 | F | 73-83 | Left MCA 11mm | MRI DSA | 1 | Intraop DSA | No complication |
| 4 | F | 52-62 | Right MCA and ACOM | MRA DSA | 2 | Intraop DSA | Transient cognitive dysfunction resolved |

ACOM = anterior communicating artery; CN = cranial nerve; CTA = computed tomography angiography; DSA = digital subtraction angiography; ICA = internal carotid artery; ICGA = indocyanine green angiography; MCA = middle cerebral artery; MRA = magnetic resonance angiography; SCA = superior cerebellar artery.

Prior to surgery, the field of view of the camera used for LSCI was co-aligned and centered with the built-in microscope camera. Additionally, the laser beam was centered with the built-in microscope camera field of view. After the craniotomy was performed, the microscope was positioned over the patient at the discretion of the neurosurgeon (R.A.). LSCI could be performed at any time when the microscope was positioned over the patient by turning on the laser illumination. LSCI did not disturb the work flow of the neurosurgeon and was performed at numerous critical times throughout the surgery. The neurosurgical co-investigator (R.A.) performed a majority of the surgeries using the surgical microscope oculars, and could observe the LSCI data in real-time on a monitor mounted next to the surgical microscope which displayed the overlayed blood flow images.

## Results

Fig. 3 shows images from single time points pre-clipping and post-clipping of the aneurysm, and during ICGA for patient 2. White light and LSCI images were acquired throughout the duration of the aneurysm clipping, and ICGA images are only available during the injection of the indocyanine green dye after the aneurysm clip was placed. In this patient, a temporary clip was placed on the patient’s carotid artery in the neck after the pre-clipping images but before the aneurysm clipping. Supplemental Video S1 shows a montage of the white light and LSCI overlay images before, during, and after the clipping procedure; Supplemental Video S2 shows a montage of the white light, LSCI overlay, and ICGA images during the injection of the ICG dye.

**Figure 3.**
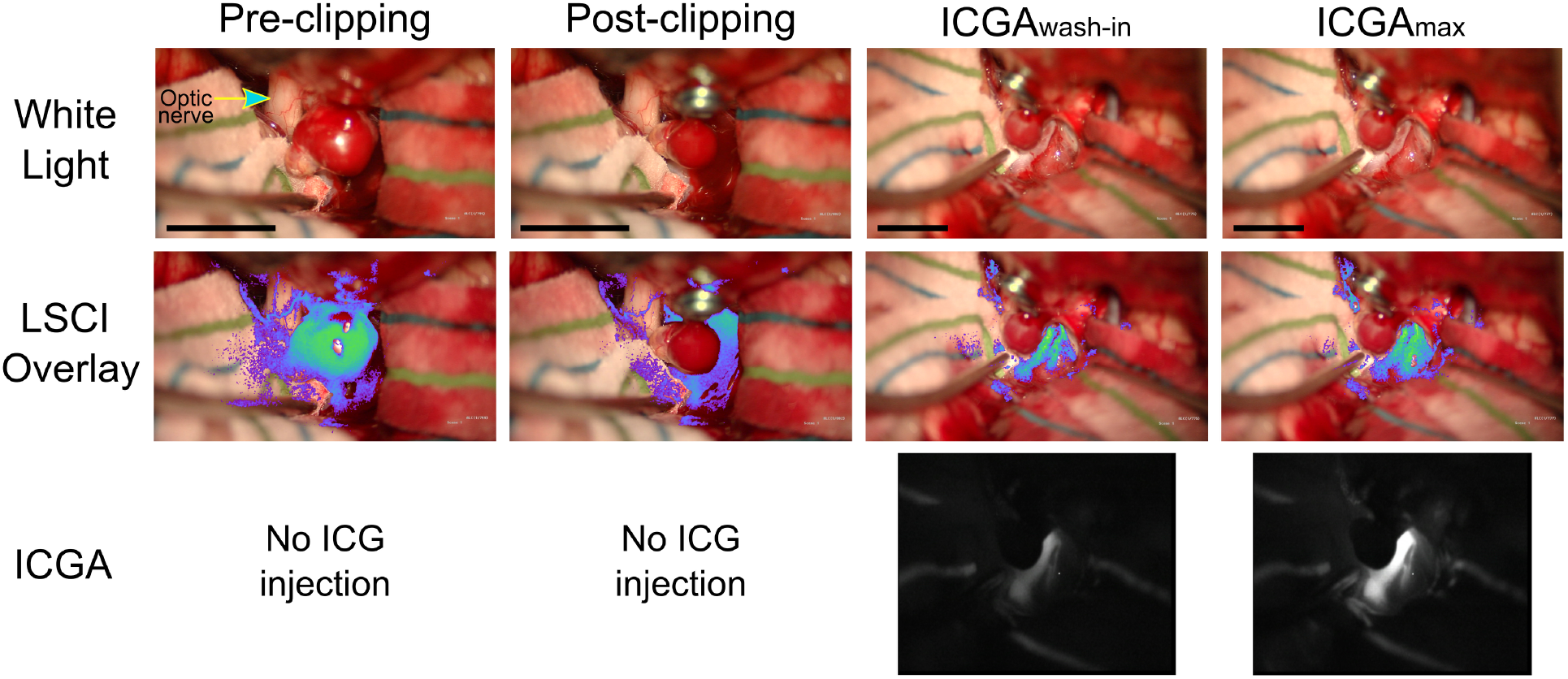
Images acquired from patient 2 before aneurysm clipping (pre-clipping), immediately after aneurysm clipping (post-clipping), and during indocyanine green angiography (ICGA) at the wash-in of the dye (ICGA_wash-in_) and at maximum fluorescence signal (ICGA_max_). Visible light images were acquired from the built-in microscope white light camera (white light); laser speckle contrast imaging (LSCI) images were acquired by an NIR-enhanced CMOS camera adapted to the microscope; and ICGA images were acquired by the built-in microscope NIR camera. LSCI overlay images were created by thresholding LSCI images and overlaying them onto the white light image with a green pseudo-color. Scale bars are 1 cm. Supplemental Video S1 shows a montage of the white light and LSCI overlay images before, during, and after the clipping procedure; Supplemental Video S2 shows a montage of the white light, LSCI overlay, and ICGA images during the injection of the ICG dye.

The pre-clipping LSCI images in Fig. 3 show there is high flow within the aneurysm prior to clipping, and that LSCI has the spatial resolution to visualize flow on the small vessels on the optic nerve (identified by arrow on pre-clipping white light image in Fig. 3). Supplemental Video S1 further illustrates that LSCI can visualize the filling of the aneurysm during cardiac cycle and the pulsatile motion of the flow within the aneurysm. This is particularly evident at a timestamp of 11 seconds in Supplemental Video S1 when the temporary clip is placed on the patient’s carotid artery in the neck causing a reduction of flow in the aneurysm. An advantage of having the LSCI instrumentation integrated into the microscope, as opposed to a stand-alone device, is that images can be acquired during operation of the microscope without interrupting the surgical work flow. This advantage is illustrated in Supplemental Video S1 at a timestamp of 59 seconds at which point the clip is placed on the aneurysm and the LSCI blood flow map immediately shows there is a cessation of flow in the aneurysm. This is similarly illustrated in the post-clipping LSCI image in Fig. 3. Approximately 5 minutes after the clipping, ICGA is used to confirm successful aneurysmal obliteration and confirm patency in surrounding vessels. In Fig. 3, the ICGA image during maximum fluorescent signal (ICGA_max_) reveals cessation of flow in the aneurysm.

The flow dynamics within the aneurysm showed in Supplemental Video S1 are quantified in Fig. 4. The pulsatile flow in the aneurysm is visible before and after the temporary clip is placed on the carotid artery in the neck. There is about a 70% reduction in average CBF after the temporary clip is placed on the carotid and the pulsatile flow within the aneurysm is clearly visible with nulls of <5% and variable peaks of 50-80% of baseline. After the clip is placed on the aneurysm, there is a significant reduction in flow and complete disappearance of pulsatile flow. The transient spikes after the clip placement are due to motion artifacts from the surgeon mechanically pushing on the aneurysm. After the microscope is repositioned, it is obvious that the CBF within the aneurysm is absent and is within the lower limit of single exposure LSCI measurements^38^ (approximately 4% of the initial CBF).

**Figure 4.**
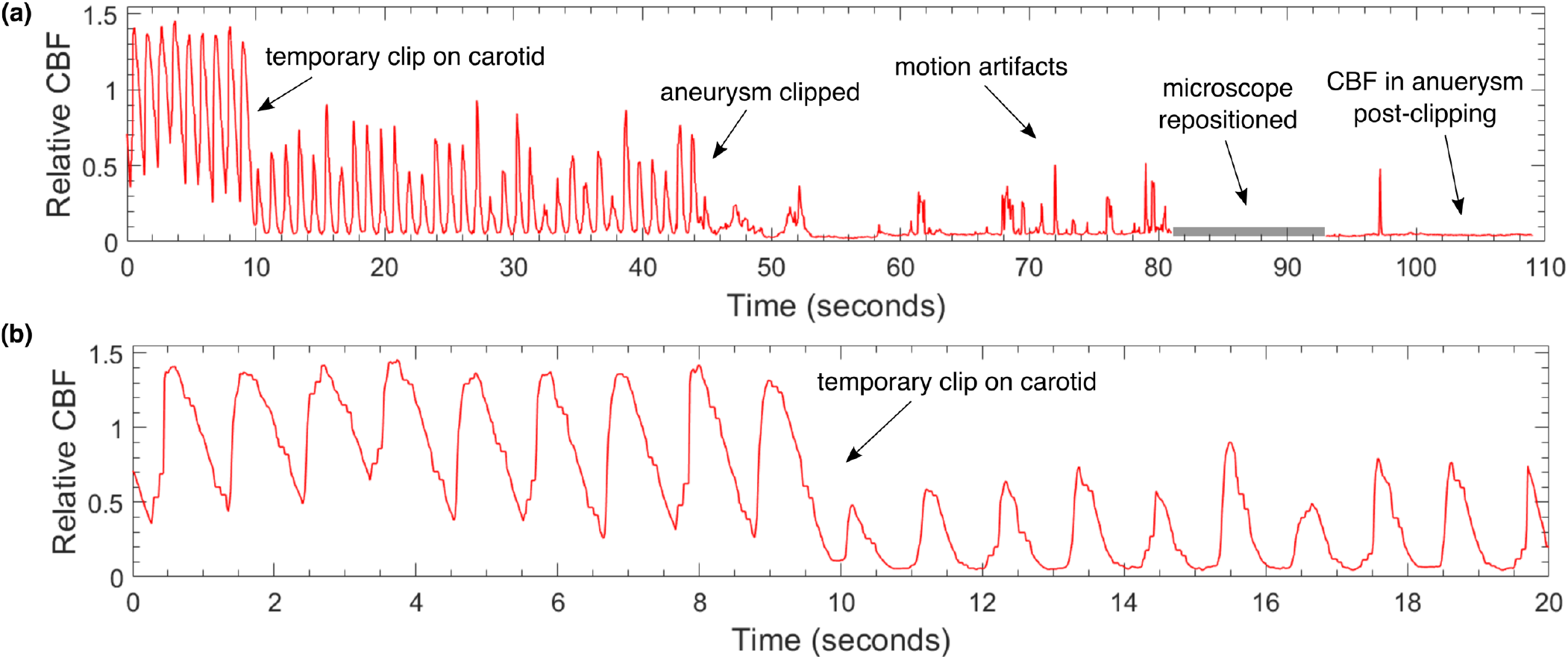
Time courses of relative cerebral blood flow (CBF) within the aneurysm from patient 2 during the clipping procedure (time matches that shown in Supplemental Video S1) for **(a)** 110 seconds and **(b)** zoomed in to the first 20 seconds. The relative CBF is normalized to the first five seconds of the data. The pulsatile nature of the flow in the aneurysm is clearly visible before and after the temporary clip is placed on the carotid artery. The reduction of CBF after the carotid temporary clip is immediately evident. Post-clipping cessation of flow is also clear. All the transients after aneurysm clipping (t=45 s) are motion artifacts.

Fig. 5 shows images from single time points during ICGA following the first aneurysm clipping procedure for patient 4, which required two clips. Supplemental Video S3 shows a montage of the white light, ICGA, and LSCI blood flow images during the ICGA procedure following the first aneurysm clipping procedure for patient 4.

**Figure 5.**
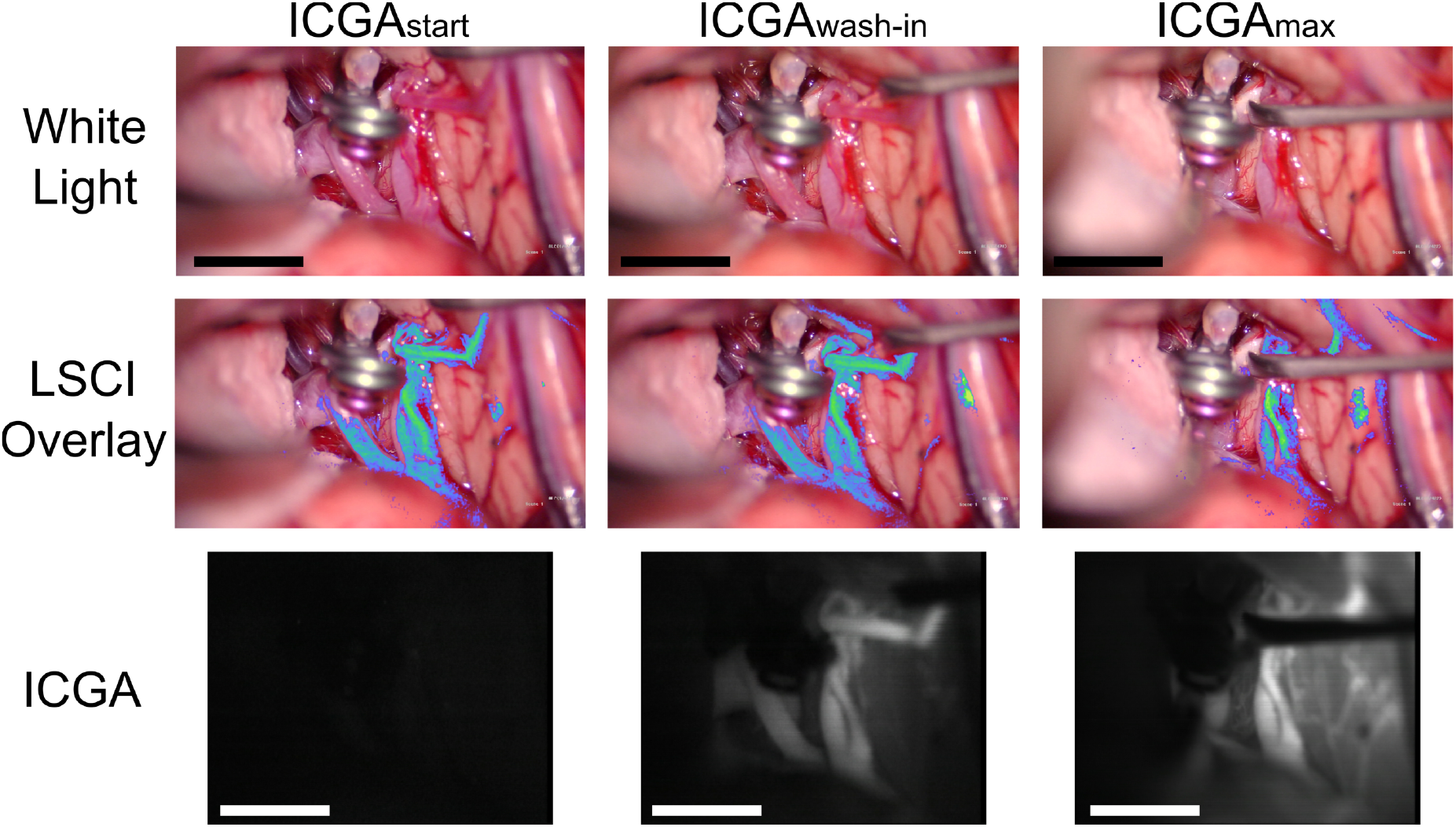
Images acquired from patient 4 during indocyanine green angiography (ICGA) at the start of the injection (ICGA_start_), wash-in of the dye (ICG_wash-in_), and at maximum fluorescence signal (ICGA_max_). Visible light images were acquired from the built-in microscope white light camera (white light); laser speckle contrast imaging (LSCI) images were acquired by an NIR-enhanced CMOS camera adapted to the microscope; and ICGA images were acquired by the built-in microscope NIR camera. LSCI overlay images were created by thresholding LSCI images and overlaying them onto the white light image. Scale bars are 1 cm.

## Discussion

Results of the current study show that LSCI can be used to continuously monitor CBF in real-time. Fig. 3 and Supplemental Video 1 demonstrate LSCI’s ability to continuously monitor CBF during critical parts of neurosurgical procedures when integrated into the surgical microscope and overlayed onto the surgeon’s view of the surgical field. LSCI reveals high flow in the aneurysm before the aneurysm clipping, and immediately after clipping, the aneurysm flow ceases while the surrounding vessels maintain perfusion. ICGA is then used to confirm patency in the surrounding vessels, but requires administration of a dye. This demonstrates that LSCI is complementary to ICGA; both can be used to determine perfusion in vessels within the surgical field of view.

Results shown in Fig. 4 demonstrate that LSCI enables quantification of flow in the aneurysm relative to a baseline value. LSCI can detect the pulsatile flow profile within the aneurysm before the aneurysm is clipped. After clipping, LSCI shows there is a >96% reduction of flow in the aneurysm relative to the initial flow and no pulsatile flow. We estimate that the uncertainty in relative flow for LSCI measurements to be approximately 5% since LSCI is sensitive to any form of motion within the tissue^38^; thus, the aneurysm may be fully occluded and yet the LSCI signal will not reduce by 100% from baseline. It is expected that the LSCI signal would still be sensitive to Brownian motion of scattering particles within the aneurysm, however this signal will be significantly smaller than flow from red blood cells into the aneurysm. While further work is needed to determine the percentage reduction of flow for a surgeon to be confident the aneurysm is fully occluded, Fig. 4 offers preliminary evidence that a 96% reduction in flow in an aneurysm as measured by LSCI indicates successful aneurysmal obliteration.

The waveforms in Fig. 4 depicting the CBF within the aneurysm match those measured with Doppler ultrasonography^39^ and a Doppler velocity wire.^40^ Fig. 4 highlights that LSCI allows for continuous CBF measurements within an aneurysm throughout neurosurgery, and thus LSCI may be useful for improving our understanding the hemodynamics in aneurysms and validating computational fluid dynamic models of the growth and rupture of aneurysms.^41,42^

Fig. 6 further illustrates the complementary nature of information obtained with LSCI and ICGA when integrated into the surgical microscope. ICGA intensity is a more direct measure of cerebral blood volume (CBV) whereas LSCI is directly sensitive to motion, and therefore is a more direct measure of CBF. ICGA wash-in can also be used to identify feeding versus draining vessels in some surgical procedures and the temporal dynamics of the fluorescence intensity can be used to estimate CBF.^43,44^ Fig. 6 shows the time-course of the fluorescence intensity change during bolus administration of ICG along with the LSCI measures of relative blood flow for the same regions of interest. In this particular example the ICG fluorescence signal saturates in the largest vessel, but it is still clear that the rise time of the ICG signal is more rapid in this vessel than the two smaller vessels, indicating higher flow. The LSCI signals from the same regions reveal similar information, but in a different manner. The LSCI signals are steady state because they are direct measures of flow, which is relatively constant over the measurement period. However, the relative flow across each region of interest is evident from the steady state values of the speckle decorrelation times. Therefore, although LSCI is unable to quantify absolute CBF, it can be used to estimate relative CBF over time or across spatial regions.

**Figure 6.**
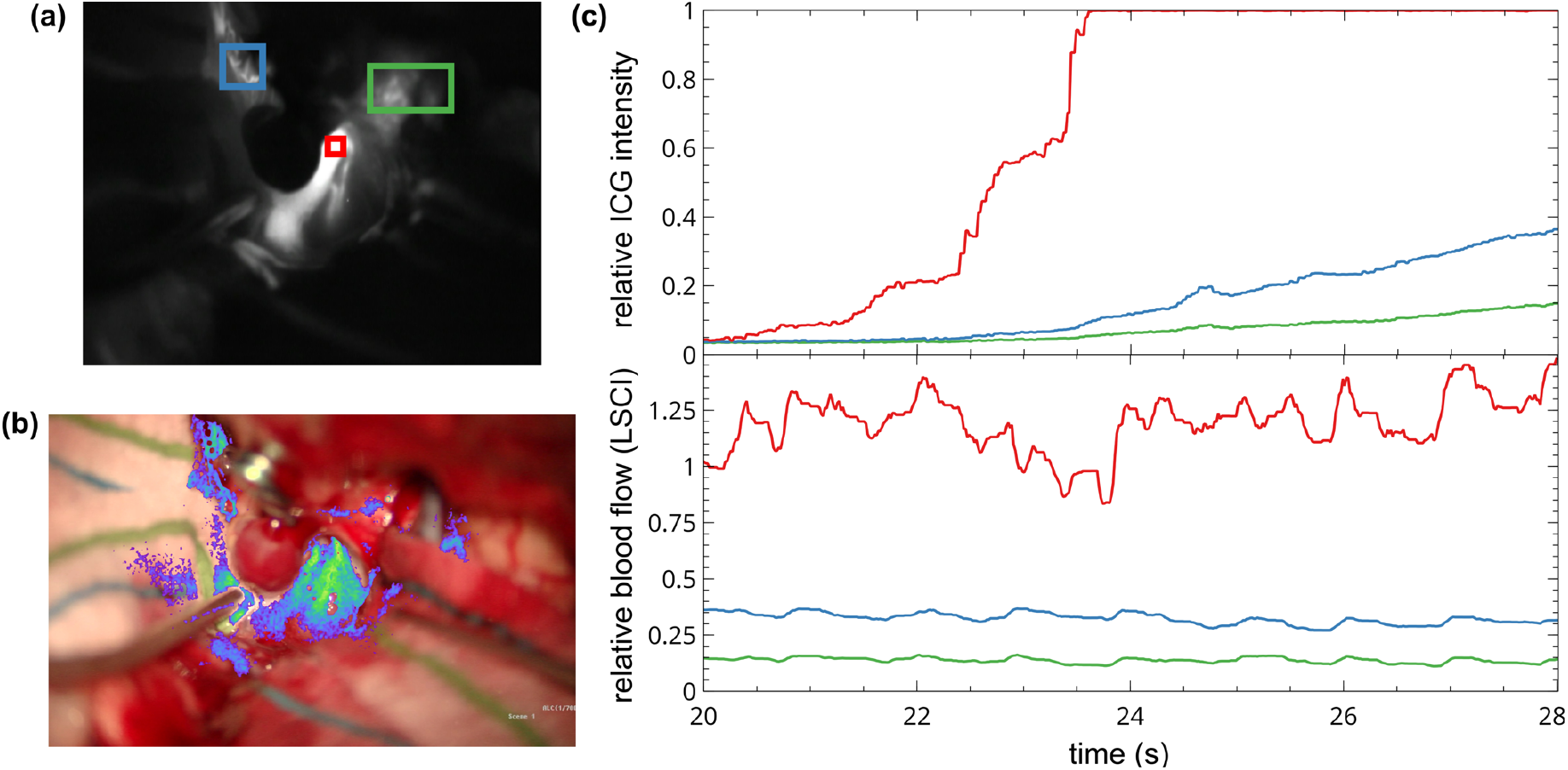
Time-course of the fluorescence intensity change during bolus administration of indocyanine green (ICG) and the laser speckle contrast imaging (LSCI) measures of relative blood flow for the same regions of interest. **(a)** ICG angiography image during wash-in of the ICG dye. Three regions of interest, for which the time-course of the fluorescence intensity is shown in (c), are highlighted in red, green, and blue. **(b)** LSCI overlay image at the same time as the ICGA image in (b). The three regions of interest selected from the LSCI overlay displayed in (c) are the same regions as highlighted in (a). **(c)** Time-course of the changes in the ICG fluorescence intensity (top) and relative blood flow measured by LSCI (bottom) during bolus administration of ICG. The three regions of interest are color coded to correspond with the respective images in (a) and (b).

Despite the differences between LSCI and ICGA, LSCI can also be used to create images that look very similar to ICGA images. Results comparing simultaneous LSCI and ICGA images shown in Fig. 7 and Supplemental Video S3 demonstrate the complementary nature of information provided by LSCI during ICGA. The LSCI and ICGA images show similar spatial information when rendered with similar green colormaps and overlayed onto the surgeon’s view.

**Figure 7.**
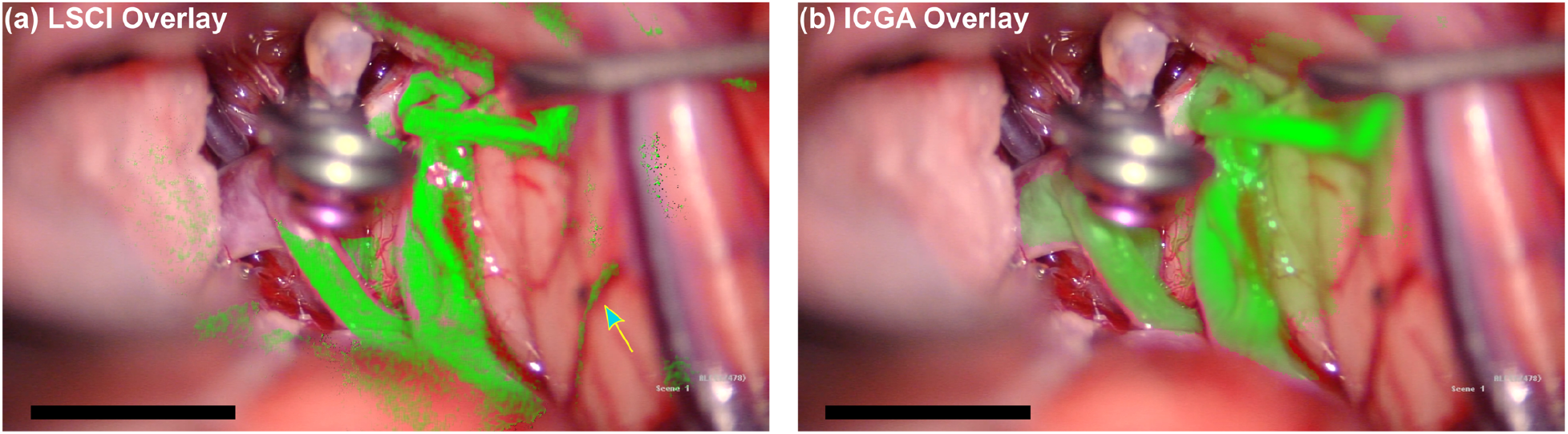
A comparison of **(a)** laser speckle contrast imaging (LSCI) overlay with **(b)** indocyanine green angiography (ICGA) overlay from patient 4. The images were created by overlaying the LSCI data and ICGA data, respectively, onto the built-in microscope white light camera and applying green-pseudo-color. The arrow in (a) highlights LSCI’s ability to detect blood flow in side wall vessels. Scale bars are 1 cm.

ICGA is better able to visualize flow in larger vessels due to ICGA using fluorescent dye as a contrast, whereas LSCI uses the inherent properties of the blood flow to scatter laser light. Conversely, LSCI has an advantage in visualizing flow in small vessels, as witnessed in Fig. 7 on the blood vessels in the side walls marked by the blue arrow; and in Fig. 3 for which LSCI shows CBF in small vessels supplying the optic nerve. ICGA also has the advantage of providing the directionality of flow during the wash-in of the dye.

One limitation of LSCI noticeable in Supplementary Videos S1-S3 is that LSCI is sensitive to motion both from blood flow and the surgeon’s mechanical force on the brain tissue. Thus, the LSCI overlay videos cannot differentiate between the two sources of motion. However, the motion induced by the surgeon can be avoided by the surgeon pausing for a moment and viewing the LSCI overlay while not pushing on the tissue. We note that the integration of the hardware for this study is not optimal, and image quality could be improved with better hardware integration.

The aim of the study was to demonstrate the potential of LSCI to monitor CBF continuously during cerebral aneurysm clipping surgery and directly compare LSCI with ICGA. The time course of relative blood flow shown in Fig. 4, and the LSCI overlay view shown in Supplementary Videos S1-S3 demonstrate the ability of LSCI to monitor CBF continuously and in real-time. The sequence of simultaneous LSCI and ICGA images shown in Figs. 3 and 5 suggest that LSCI and ICGA provide complementary information about CBF as both can be used to determine perfusion in a vessel.

## Conclusions

Our results suggest that LSCI can provide continuous and real-time CBF visualization without affecting the surgeon work-flow or requiring a contrast agent, and thus is a promising tool for continuous CBF monitoring during surgery. We performed LSCI of CBF at critical parts of aneurysm clipping neurosurgery, demonstrating that LSCI and ICGA are complementary tools for visualizing CBF to aid surgical decision making. LSCI was performed continuously, whereas ICGA had a refractory period and was limited to one or two time points.

## Data Availability

De-identified data upon request.

## Acknowledgements

This work was funded by a grant from the National Institutes of Health (EB011556). The authors thank the neurosurgical staff at the Dell Seton Medical Center, particularly John Hourie and Fred Canales. We are grateful to Tina Adrean, Cierra Grubbs, Emily Niewiarowski, Dayal Rajagopalan, and Akhil Surapaneni for assistance in consenting patients. We are appreciative for help from Seth Sork and Brian Schoelpple in configuring the microscope.

## Author Contributions

D.R.M, R.A., and A.K.D. conceived of the study and design. D.R.M., R.A., and A.K.D. acquired the data. D.R.M, C.T.S., and A.K.D. performed post-imaging analysis and interpretation of LSCI and ICGA data; R.A. performed interpretation of ICGA, CTA, and DSA data. D.R.M., R.A., and A.K.D. drafted the manuscript. D.R.M. and C.T.S. finalized the main figures and supplementary videos. All authors provided critical revisions.

